# Students Perceptions of an innovative and resilient approach in teaching human anatomy without cadaveric resources: the case of the Medical School of the University of Burundi

**DOI:** 10.64898/2026.02.21.26346765

**Authors:** C.P. Baramburiye, D. Kamatari, J.C. Mbonicura, F. Nduwimana, P. Hakizimana, L. Ndayisaba, G. Ndayizeye, P. Banderembako

**Author notes:** **Corresponding author:** Clovis Paulin BARAMBURIYE, MD, FCS (ECSA).

## Abstract

**Background:** At the school of medicine of the University of Burundi, we have faced the challenge of lack of dissection facilities and cadaveric resources, and hence, we tried to use evidence-based alternative teaching methods to meet the student needs and enhance learning outcomes in anatomy. The aim of this study was to collect students’ perceptions regarding this innovative teaching method of anatomy and share our experience to professionals working in similar environments.

**Methods:** We have designed a multimodal approach where first year medical students were first exposed to the topics during lectures and practical sessions were held afterwards in small groups using four materials including: YouTube dissection videos, 3d plastic models, anatomy drawings and the 3D4Medical virtual anatomy app. A Likert scale questionnaire including questions regarding the perceived achievement of learning objectives, 3d understanding of the structures and engagement with the content was distributed to them. Moreover, we have performed paired samples contingency tables and McNemar tests to check the statistical significance of the results in comparing the different didactic methods.

**Results:** The majority of the students had positive perceptions regarding the multimodal approach. They preferred the combination of lectures and practicals rather than these didactic methods used separately (p<0.001). Furthermore, regarding the tools used in the practical sessions, the combination of the tools was also significantly preferred (p<0.001). The virtual anatomy app was significantly superior to the YouTube dissection videos in perceived achievement of 3d understanding (p=0.018). Moreover, the students agreed that being taught anatomy by surgeons has helped in bringing in more context that is useful to transfer the learnt knowledge into real life situations (p<0.001).

**Conclusion:** Efficient teaching of anatomy without cadaveric materials can be achieved by combining multiple didactic tools that promote active learning and enhance 3d understanding of the material.

## Introduction

The mandatory use of cadaveric materials in effective teaching of anatomy has been an active point of discussion in medical education literature. It has been argued that early exposure of students to cadaveric materials even though they are not actively dissecting the specimen, may increase the values of compassion and respect to the human subject, which will be part of the cores of their careers as medical providers. ^[1, 2]^ On another hand, it is has been argued that undergraduate students do not have to use cadaveric materials to efficiently learn anatomy and that it should be reserved for graduate students who are pursuing surgical careers. ^[3]^ Moreover, the old fashion didactic dissection-based teaching model has gradually given place to a more multimodal, student-centered approach. A combination of physical and virtual models has been agreed on by many research reports; these models include plastic models, virtual models such as the Anatomage table, 3d apps and YouTube videos. Furthermore, students-centered approaches have been promoted such as team-based learning and peer teaching. ^[4], [5], [6], [7]^

Teaching anatomy in a low resource setting especially without cadaveric material, has been directed towards the use of the multimodal approach with alternative tools. ^[8]^ At the faculty of medicine of the University of Burundi, we have faced this challenge of lack of dissection facilities and cadaveric resources and tried to use evidence-based alternative teaching methods to meet the student needs and enhance learning outcomes in anatomy. We have designed a multimodal approach where first year medical students were first exposed to the topics during lectures, afterwards we have organised practical sessions in small groups using four materials including YouTube dissection videos, plastic models, anatomy drawings and the 3D4Medical virtual anatomy app.

## Methods

The study included first year medical students from the 2024/2025 batch of the University of Burundi. Upon completion of the visceral and musculoskeletal anatomy modules where we have used the multimodal teaching approach; we have organised a feedback session to collect their perceptions regarding their learning experience. For this purpose, we have used a questionnaire that included questions regarding their favorite didactic methods, and their perceived achievement of learning objectives, especially 3d understanding and engagement with the teaching content using a Likert scale. Responses of the those who have given informed consent where considered for analysis and the submission of answers was anonymous.

## Results

A total of 102 participants were present during the feedback session, among which 100 consented on the questionnaire to participate in the study. According to the sample size we calculated using Slovin’s formula, we needed 94 participants to have significant results representative of the batch population which was of 123 students.

n= N/1+Nd^2^

n: sample size,

N: Population = 123 students

d: margin of error = 0,05

Regarding the favorite learning method, none of the participants selected lectures only as they favorite didactic method; 79% favored a combination of lectures and practical sessions while 21% favored practical sessions only. Moreover, for the practical sessions, the combination of all the tools (YouTube dissection videos, image annotations, virtual anatomy app and the plastic models) was the most favorite practical scheme for 67% of the participants. We have done comparison tests using McNemar test between the tools used in practical sessions 2 by 2 and discovered that the combination of them was significantly favored compared to each single tool (p<0.001). (**Table 1)**

**Table 1:** Favored practical scheme.

| **Table 1: Favored practical scheme** |  |  |  |  |  |
| --- | --- | --- | --- | --- | --- |
| Binomial Test |  |  |  |  |  |
|  | Level | Count | Total | Proportion | p |
| Favors Virtual anatomy | No | 86 | 100 | 0.860 | <.001 |
|  | Yes | 14 | 100 | 0.140 | 1.000 |
| Favors plastic models | No | 91 | 100 | 0.910 | <.001 |
|  | Yes | 9 | 100 | 0.090 | 1.000 |
| Favors YouTube dissection videos | No | 96 | 100 | 0.960 | <.001 |
|  | Yes | 4 | 100 | 0.040 | 1.000 |
| Favors Image annotation | No | 94 | 100 | 0.940 | <.001 |
|  | Yes | 6 | 100 | 0.060 | 1.000 |
| Favors Combination (2) | No | 33 | 100 | 0.330 | 1.000 |
|  | Yes | 67 | 100 | 0.670 | <.001 |

Note. $H_a$ is proportion > 0.5

Furthermore, the virtual anatomy app was significantly more preferred when compared the use of YouTube dissection videos for practical sessions (p=0.018). (**Table 2)**

**Table 2:** Comparison of YouTube dissection videos and Virtual anatomy app.

|  |  |  |  |
| --- | --- | --- | --- |
| Contingency Tables |  |  |  |
| Favors Virtual anatomy | Favors YouTube dissection videos |  | Total |
|  | No | Yes |  |
| No | 82 | 4 | 86 |
| Yes | 14 | 0 | 14 |
| Total | 96 | 4 | 100 |

| McNemar Test |  |  |  |
| --- | --- | --- | --- |
|  | Value | df | p |
| $\chi^2$ | 5.56 | 1 | 0.018 |
| N | 100 |  |  |

The YouTube videos, 3D4medicals virtual anatomy app and the plastic models have all received a majority of positive perceptions regarding the enhancement of the 3d understanding of the structures. (**Table 3**,**4**,**5)** Moreover, 75% of the participants disagreed that only lectures could make them achieve learning objectives. Also, 92% of the participants agreed that small groups disposition in the practical sessions helped them to engage more with the material and hence understand better the teaching materials. The fact of being taught by surgeons was perceived to increase clinical context by 90% of the participants. Finally, most of the participants (91%) agreed that the access to cadaveric teaching material could have increased even more their understanding of the topics.

**Table 3:** Perceived enhancement of 3d understanding with YouTube videos.

| Levels | Counts | % of Total | Cumulative % |
| --- | --- | --- | --- |
| Agree | 49 | 49.0 % | 49.0 % |
| Disagree | 6 | 6.0 % | 55.0 % |
| Neutral | 12 | 12.0 % | 67.0 % |
| Strongly agree | 25 | 25.0 % | 92.0 % |
| Strongly disagree | 8 | 8.0 % | 100.0 % |

**Table 4:** Perceived enhancement of 3d understanding with Virtual Anatomy app.

| Levels | Counts | % of Total | Cumulative % |
| --- | --- | --- | --- |
| Agree | 45 | 45.0 % | 45.0 % |
| Disagree | 3 | 3.0 % | 48.0 % |
| Neutral | 1 | 1.0 % | 49.0 % |
| Strongly agree | 48 | 48.0 % | 97.0 % |
| Strongly disagree | 3 | 3.0 % | 100.0 % |

**Table 5:** Perceived enhancement of 3d understanding with Plastic models.

| Levels | Counts | % of Total | Cumulative % |
| --- | --- | --- | --- |
| Agree | 53 | 53.0 % | 53.0 % |
| Disagree | 8 | 8.0 % | 61.0 % |
| Neutral | 4 | 4.0 % | 65.0 % |
| Strongly agree | 31 | 31.0 % | 96.0 % |
| Strongly disagree | 4 | 4.0 % | 100.0 % |

## Discussion

Teaching anatomy in low resources can be challenging especially when cadaveric dissection resources are lacking. Efforts have been made to find alternative solutions which could allow at some extent acceptable learning outcomes. ^[8]^ The paradigm of a multimodal approach in teaching anatomy is not a new concept and has been utilized in medical schools around the world. The combination of didactic tools rather than the sole use of cadaveric dissection was proved to be the best approach in achieving learning outcomes. ^[4]^ This fact is confirmed in our results when the students agree that they prefer the combination of teaching methods rather than just one.

Moreover, the importance of three-dimensional models, whether physical or virtual, in enhancing visuo-spatial ability and hence improving the understanding of the anatomical structures is a well reported fact ^[6]^, this is also shown in our results where students preferred more 3 dimensional models. When comparing the virtual models, we discovered that the virtual anatomy app was preferred to YouTube dissection videos and the reason for that could be that the use of the app was more interactive which allowed the students to view in detail any given structure at their own pace, which we believe rendered the learning process more engaging and hence more interesting.

Furthermore, it is thought that the use of active teaching methods increases the student engagement with the teaching materials and understanding of the subjects ^[9]^; our students confirmed this fact as they preferred small group interactive sessions rather than lectures. Adding clinical context in the basic science is crucial for the students to be able to transfer the learnt knowledge in the classroom later on during clinical clerkships, one of the ways to do so is having clinicians teaching basic science subjects which includes involving surgeons teaching anatomy. ^[10, 11]^ The latter has been confirmed by the students that perceived an added value of having surgeons teaching them anatomy since they were correlating the subjects to real life clinical situations.

Although students had positive perceptions regarding our innovative teaching method, the majority have agreed that they think adding cadaveric materials in the didactic tools would have an added positive impact in their learning process. Although some has argued that undergraduate students do not benefit much from cadaveric dissection but that is rather a tool suited for students that have chosen surgical careers, others support that early exposure to cadaveric dissection will enhance 3-d understanding of the structures, distinguish the texture of the different tissues of the human body and promote of respect to the human body which are crucial assets in the clinical career. ^[3] [2] [8]^ Hence, although we have developed a method that would help the students achieving positive learning outcomes without cadaveric tools, it is due to a low resource setting, however, we join the opinions that students can benefit even more if cadaveric dissection is included when resources will allow it.

## Conclusion

Efficient teaching of anatomy without cadaveric materials can be achieved by combining multiple didactic tools that promote active learning and enhance 3d understanding of the material. However, including cadaveric dissection is a good bonus that one should use if resources allow it.

## Data Availability

All data produced in the present study are available upon reasonable request to the authors

## Ethical considerations

The conduct of this study has been approved by institutional review board of our school of medicine.

## Acknowledgements

To the medical students who have accepted to willingly participate in this study.

## Conflict of interest

The authors declare that there is no conflict of interest related to this study.

## Funding sources

None

## Notes

### Competing Interest Statement

The authors have declared no competing interest.

### Funding Statement

This study did not receive any funding

### Author Declarations

Ethics commitee of University of Burundi's Faculty of Medicine gave ethical approval for this work

